# Metagenomic analysis during capecitabine therapy reveals microbial chemoprotective mechanisms and predicts drug toxicity in colorectal cancer patients

**DOI:** 10.1101/2024.10.11.24315249

**Authors:** Lars E. Hillege, Kai R. Trepka, Janine Ziemons, Romy Aarnoutse, Benjamin G. H. Guthrie, Judith de Vos-Geelen, Liselot Valkenburg-van Iersel, Irene E.G. van Hellemond, Arnold Baars, Johanna H.M.J. Vestjens, John Penders, Adam Deutschbauer, Chloe E. Atreya, Wesley A. Kidder, Peter J. Turnbaugh, Marjolein L. Smidt

## Abstract

**Purpose:** Unpredictable chemotherapy side effects are a major barrier to successful treatment. Cell culture and mouse experiments indicate that the gut microbiota is influenced by and influences anti-cancer drugs. However, metagenomic data from patients paired to careful side effect monitoring remains limited. Herein, we focus on the oral fluoropyrimidine capecitabine (CAP). We investigate CAP-microbiome interactions through metagenomic sequencing of longitudinal stool sampling from a cohort of advanced colorectal cancer (CRC) patients.

**Methods:** We established a prospective cohort study including 56 patients with advanced CRC treated with CAP monotherapy across 4 centers in the Netherlands. Stool samples and clinical questionnaires were collected at baseline, during cycle 3, and post-treatment. Metagenomic sequencing to assess microbial community structure and gene abundance was paired with transposon mutagenesis, targeted gene deletion, and media supplementation experiments. An independent US cohort was used for model validation.

**Results:** CAP treatment significantly altered gut microbial composition and pathway abundance, enriching for menaquinol (vitamin K2) biosynthesis genes. Transposon library screens, targeted gene deletions, and media supplementation confirmed that menaquinol biosynthesis protects *Escherichia coli* from drug toxicity. Microbial menaquinol biosynthesis genes were associated with decreased peripheral sensory neuropathy. Machine learning models trained in this cohort predicted hand-foot syndrome and dose reductions in an independent cohort.

**Conclusion:** These results suggest treatment-associated increases in microbial vitamin biosynthesis serve a chemoprotective role for bacterial and host cells, with implications for toxicities outside the gastrointestinal tract. We provide a *proof-of-concept* for the use of microbiome profiling and machine learning to predict drug toxicities across independent cohorts. These observations provide a foundation for future human intervention studies, more in-depth mechanistic dissection in preclinical models, and extension to other cancer treatments.

## Introduction

With the emerging field of pharmacomicrobiomics, it has become increasingly evident that bi-directional interactions exist between the gut microbiota and numerous drugs, including those not traditionally classified as antibiotics^1, 2^. As such, the gut microbiota is both affected by chemotherapy and may alter chemotherapy outcomes. Treatment-related toxicity considerably influences the quality of life of patients with colorectal cancer (CRC), often causing treatment delays and dose reductions that impact efficacy^3^. Therefore, understanding the role of the gut microbiota in chemotherapy is of high clinical importance.

Capecitabine (CAP) is a commonly used chemotherapy in patients with CRC, either as monotherapy or with other agents^4, 5^. CAP is administered as an oral prodrug and is sequentially converted by host enzymes into the active compound 5-fluorouracil (5-FU), which exerts its anticancer effects by disrupting DNA synthesis and RNA processing^6^. Subsequently, 5-FU is metabolized into the inactive metabolite dihydrofluorouracil by host clearance enzyme dihydropyrimidine dehydrogenase^6^.

CAP has two major drawbacks: limited response rates and toxicity. While there have been significant advancements in the early-stage CRC setting, overall response rates in the advanced CRC setting remain modest, falling between 34-42%^7, 8^. Further, many patients suffer from CAP-induced toxicity, with up to 57% requiring dose alterations or treatment discontinuation^9^. Common side effects include diarrhea, hand-foot-syndrome (HFS), and oral mucositis^7, 10–12^.

The gut microbiota modulates CAP efficacy and toxicity in mouse models. Specific gut bacteria harbor a bacterial homologue of host DPYD, encoded by the *preTA* operon^13, 14^. *preTA*-containing bacteria can metabolize and inactivate 5-FU, modulating treatment efficacy and toxicity in mice^13, 15^. Beyond direct drug metabolism, gut *Lactobacillus* potentiate the anti-tumor effects of CAP through immunologic and pro-apoptotic effects^16, 17^.

In a clinical setting, we detected only slight CAP-induced bacterial shifts in our prior 33 patient cohort with advanced CRC using 16S rRNA sequencing^11, 15^. Fecal levels of microbially-derived valerate and caproate decreased significantly during CAP treatment, and baseline levels of the iso-butyrate were associated with tumor response^18^. Taken together, these studies highlighted (1) the need to measure gut microbial functional potential; (2) the importance of mechanistic follow-up; and (3) the utility of validation on a separate cohort. Therefore, the current study investigated CAP-microbiome interactions by performing metagenomic sequencing of stool samples from a larger cohort of advanced CRC patients with detailed toxicity data.

## Methods

### 1. Study design and population

This prospective longitudinal cohort study was conducted in Maastricht University Medical Center (MUMC+), Catharina Hospital Eindhoven, Hospital Gelderse Vallei and VieCuri Medical Center in the Netherlands, following the previously published study protocol (NL-OMON29314/NTR6957)^19^. This study was approved by the Medical Ethics Committee azM/UM (METC 16-4-234.1) and conducted in accordance with the Declaration of Helsinki and Good Clinical Practice. Each patient provided written informed consent. Patients were eligible if diagnosed with metastatic or unresectable CRC with planned CAP treatment, with or without the VEGF inhibitor bevacizumab. Exclusion criteria included radiotherapy within two weeks of enrollment, other systemic therapy within one month of inclusion, antibiotic use within three months of enrollment, microsatellite instability (MSI), impaired renal function (creatinine clearance <30 ml/min), and (sub)total colectomy and/or ileostomy. CAP therapy was administered in a 3-week cycle, consisting of two weeks of oral CAP at a dose of 1000-1250 mg/m² (twice daily), followed by a one-week rest period. Treatment was adjusted if deemed necessary by the treating oncologist.

### 2. Sample and data collection

#### 2.1 Fecal samples

Fecal samples were collected at three timepoints: before CAP initiation (t_1_), during week 2 of CAP cycle 3 (t_2_), and after week 3 of cycle 3 (t_3_). Fecal samples were collected by patients at home in preservation-free tubes (Sarstedt) and stored in freezers. Frozen samples were transported to the hospital in cooled containers (Sarstedt) and stored at −80°C long-term.

#### 2.2 Clinical data and chemotherapy-induced toxicity

Patients completed questionnaires on health-related characteristics and medical history. Chemotherapy-induced toxicities were self-reported by the patients and scored based on the Common Terminology Criteria for Adverse Events (CTCAE v4.0)^20^. The questionnaire encompassed nausea (0-3), vomiting (0-5), diarrhea (0-5), unintended weight loss (0-3) constipation (0-5), peripheral sensory neuropathy (PSN) (0-5), oral mucositis (0-5), HFS (0-3), fever (0-5), alopecia (0-2), and fatigue (0-3). Additional data about medical history, tumor characteristics, medications, surgery, dihydropyrimidine dehydrogenase (DPYD) deficiency, and CAP dose adjustments were collected from medical records.

### 3. Gut microbiome analysis

ZymoBIOMICs 96 MagBead DNA Kit was used for fecal DNA extractions (156 samples from 56 patients), with extraction, library preparation, sequencing, and read mapping performed as described previously^15^. Taxa abundances were central log ratio (CLR)-transformed. Pathway/gene abundances were normalized to reads per kilobase per genome equivalent using microbeCensus^21^. Shannon diversity was calculated using vegan command diversity^22^. PERMANOVA was performed using vegan commands vegdist (CLR-Euclidean/Aitchison distance) and adonis2^22^ to compare patient demographics and patient-reported toxicities (any grade) to the baseline microbiome. Differential abundance was calculated using linear mixed effects modeling with time as a fixed effect and patient as a random effect (nlme command lme), followed by False Discovery Rate (FDR) correction. Phylogenetic trees were obtained by pruning the MetaPhlAn 4 tree^23^ and visualized using ggtree^24^.

### 4. In vitro studies of fluoropyrimidine toxicity in E. coli

#### 4.1 Transposon sequencing experiment

We performed *E. coli* transposon mutant fitness assays as described previously^25^. A thawed transposon library aliquot was grown overnight in 25 mL Luria broth (LB) with 50 μg/mL kanamycin at 37°C with 225 rpm shaking. Cells were then inoculated into competitive growth assays in fluoropyrimidines (500 μM CAP, 5-fluorouracil, 5’deoxy-5-fluorocytidine) or vehicle. Assays were performed in duplicate in 200 μL M9 minimal media with starting OD_600_=0.02. After 48 hours, cell pellets were collected and gDNA extracted with ZymoBIOMICS 96 MagBead DNA kit (ZymoResearch D4302) per manufacturer protocol. We performed barcode sequencing as previously described, averaging independent insertions at the gene level and calculating log-ratios^26^. A quantile-quantile method was used to determine significance (abs(ln(FC))>0.25, abs(log(VehFitness))<0.05, **Supplemental Figure 3a**). Overlap between conditions was visualized using UpSetR^27^. Gene set enrichment analysis was performed using clusterProfiler function enrichKEGG (universe=library, organism=“eco”, pvalueCutoff=0.01)^28^.

#### 4.2 5-FU sensitivity experiments

*E. coli* BW25113 wild-type and *ΔmenF::Kan^R^* were obtained from the Keio collection^29^ and streaked on LB with 30 μg/mL kanamycin. Colonies were subcultured overnight in Brain Heart Infusion (BHI) in an anaerobic chamber (Coy Laboratory Products) at 37°C with an atmosphere of 3% H_2_, 20% CO_2_, and balance N_2_. 5-FU (MilliporeSigma 343922) was assayed at 0 and 500 μM. Vitamin K2 (MilliporeSigma V9378) was dissolved in methanol, supplemented 1% (v/v), and assayed at 0 and 0.1 μg/ml. Uracil (MilliporeSigma U0750) was assayed at 0 and 50 μM. 3 µL seed culture diluted to OD_600_=0.1 was inoculated with 197 µL media±drug in a 96-well plate. Plates were covered (Breathe-Easy sealing membrane) and incubated anaerobically at 37°C for 24 hr in a plate reader (Biotek Gen5), with 1 min linear shake prior to OD_600_ readings every 15 min. Carrying capacity was determined using package Growthcurver^30^.

### 5. Random forest modeling

Two cohorts were used for modeling: a training cohort (this Netherlands cohort, 48 patients with baseline stool sequencing and cycle 3 toxicity data), and an independent validation cohort (U.S. cohort, *n* = 38 patients with baseline stool sequencing and on-treatment toxicity data)^15^. HFS and dose adjustment were selected as targets due to available toxicity data in both cohorts, while PSN was chosen due to the strong microbiome signal in our analysis. Features (CLR-normalized KOs) were selected by applying the Boruta algorithm^31^ to the training cohort for HFS and dose adjustments. For PSN, the top 10 pathways from differential abundance testing were used as features, with no external validation cohort (no PSN data for U.S. cohort). A random forest model was fitted using these features (500 trees, leave-one-out-cross validation (LOOCV)) using packages randomForest^32^ and caret^33^. Within-cohort model accuracy was evaluated by training 100 separate models, validating on left-out samples (LOOCV), and plotting the mean and 95% confidence interval of receiver operating curves using pROC^34^. For HFS and dose adjustments, model generalizability was validated in the U.S. cohort.

### 6. Statistical analysis

Statistical analysis was performed in R (v4.2.1)^35^, with plots generated using ggplot2 (v3.5.1) and ggpubr (v0.6.0)^36, 37^. Statistical tests are specified in the text/figure legends where used and summarized here. PERMANOVA (CLR-Euclidean ordination) was used to test compositional differences in taxa vs patient characteristics, and gene pathways vs toxicity. Linear mixed-effects modeling (time as a fixed effect, patient as a random effect) was used to identify time-dependent changes in taxa/genes. T-tests, Spearman’s correlation, and likelihood-ratio tests were used to identify relationships between categorical/continuous, continuous/continuous, and categorical/categorical variables, respectively. Significance was determined as *p*<0.05 (individual tests) or Benjamini-Hochberg FDR<0.2 (multiple hypothesis correction).

## Results

In total, 56 patients were enrolled (**Table 1**). 71% were treated with bevacizumab in combination therapy, 70% had left-sided tumors, 29% had a colostomy, and 55% had received prior systemic treatment, mainly CAP with oxaliplatin (CAPOX; **Table 1**). The majority (79%) of the patients had previously undergone surgical resection of their primary tumor, including 18 with rectum resection, 9 with sigmoid resection, 10 with right hemicolectomy, 4 with left hemicolectomy, and 1 with extended left hemicolectomy (**Supplemental Table 1**).

**Table 1.** Baseline characteristics.

| **Clinical characteristics (n=56)** |  |
| --- | --- |
| Age (years) - Mean (SD) | 72.9 (7.6) |
| Gender - N (%) |  |
| Male | 36 (64.3) |
| Female | 20 (35.7) |
| BMI (kg/m<sup>2</sup>) - Mean (SD) | 26.77 (4.4) |
| Co-treatment with bevacizumab - N (%) | 40 (71.4) |
| Tumor sidedness - N (%) |  |
| Left-sided (descending colon, sigmoid colon, rectum) | 39 (69.6) |
| Right-sided (cecum, ascending colon, transverse colon) | 16 (28.6) |
| Missing data | 1 (1.8) |
| Colostomy in situ - N (%) | 16 (28.6) |
| **Prior treatments and medication (n=56)** |  |
| Prior systemic treatment (>1 month before inclusion)* - N (%) | 31 (55.4) |
| CAP (with or without B) | 6 (10.7) |
| CAP+RT | 11 (19.6) |
| CAPOX (with or without B) | 24 (42.9) |
| CAPIRI+P | 1 (1.8) |
| FOLFIRI | 2 (3.6) |
| FOLFIRINOX (with or without B) | 2 (3.6) |
| TAS+B | 1 (1.8) |
| Prior chemoradiation (>1 month before inclusion) - N (%) | 11 (19.7) |
| Antibiotic use last year (>3 months before inclusion) - N (%) | 23 (41.1) |
| Colorectal surgery in the past - N (%) | 44 (78.6) |
| Proton pump inhibitor use - N (%) | 18 (32.1) |

\*CAP: Capecitabine; B: Bevacizumab; RT: Radiotherapy; CAPOX: Capecitabine + Oxaliplatin; CAPIRI: Capecitabine + Irinotecan; P: Pembrolizumab; FOLFIRI: 5-Fluorouracil + Irinotecan; FOLFIRINOX: 5-Fluorouracil + Irinotecan + Oxaliplatin; TAS: Trifluridine and Tipiracil. Each count represents a single patient - if a patient had multiple previous rounds of a single treatment (i.e. CAPOX), this is still only counted as 1 event. Total percentage exceeds 100 because some patients had multiple distinct prior treatments.

A total of 157 stool samples were collected across the 3 timepoints (2-3 stool samples/subject; **Figure 1a**). DNA was extracted and used for deep metagenomic sequencing, resulting in 39.9±2.5 million high-quality sequencing reads/sample (11.7±0.7 Gbp, **Supplemental Table 1**). Inter-individual differences in microbial community accounted for 83% of the variation in the combined dataset, as evidenced by species-level principal coordinate analysis (**Supplemental Figure 1a**).

**Figure 1.**
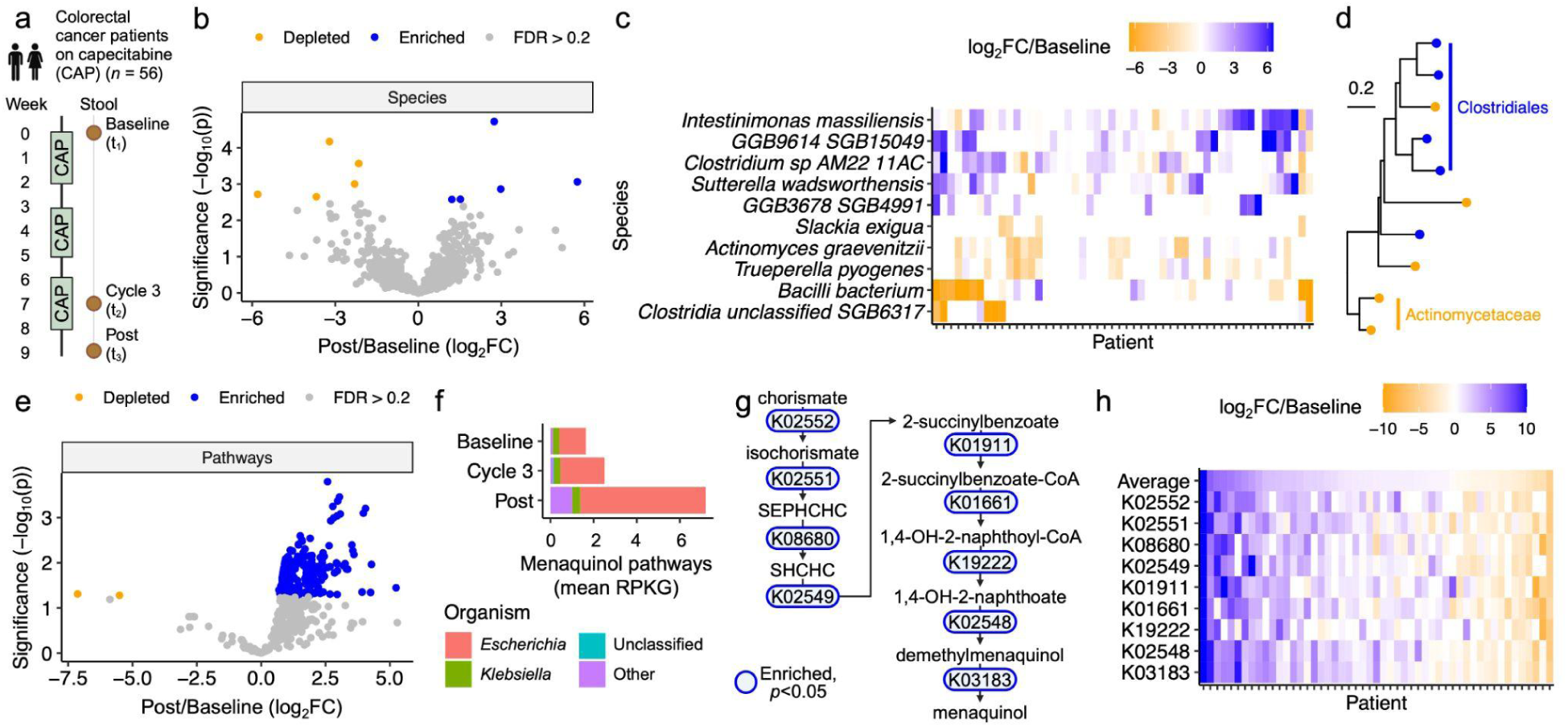
Capecitabine (CAP) alters the human gut microbiome. **(a)** Study design. Patients with advanced colorectal cancer (CRC) were treated with three cycles of CAP, with stool collected at baseline (t_1_), during cycle 3 (t_2_), and post-treatment (t_3_). Created with BioRender.com. **(b)** Volcano plot of species post-treatment (t_3_) vs baseline (t_1_). Points represent significantly enriched (blue) and depleted (orange) species (FDR<0.2). **(c)** Heatmap of differentially abundant species from **(b)**, with patients and species ordered by McQuitty hierarchical clustering of log_2_ fold change (log_2_FC) of post (t_3_) vs baseline (t_1_). **(d)** Phylogenetic tree of differentially abundant species from **(b)**, with labels for clades where treatment affected multiple clade members similarly [enriched (blue) or depleted (orange)]. **(e)** Volcano plot of HUMAnN 3.0 gene pathways at post (t_3_) vs baseline (t_1_). Points represent significantly enriched (blue) and depleted (orange) pathways (FDR<0.2). 7 of the top 10 most significantly altered pathways are menaquinol biosynthesis pathways. **(f)** Genera of microbes contributing to menaquinol biosynthesis pathways. **(g)** KOs shared across all enriched menaquinol biosynthesis pathways in **(f)**. Blue indicates *p*<0.05. **(h)** Heatmap of all KOs from **(g)**, with patients ordered by average log_2_FC (top row, “Average”) and KOs ordered by occurrence in the menaquinol biosynthesis pathway. **(b,e,g)**: *p*-value: mixed effects model of central log ratio (CLR)-normalized abundance vs time, with patient as a random effect.

Multiple patient characteristics were associated with variations in baseline microbial diversity and taxonomic composition. We binarized all baseline characteristics and performed t-tests to identify significant associations with the Shannon diversity index (**Supplemental Figure 1b**). Antibiotic use within the past year (>3 months before inclusion) and prior systemic treatment (>1 month before inclusion) were both associated with significantly lower baseline diversity, with most of the signal coming from patients who had both prior systemic treatment and prior antibiotic use (**Supplemental Figures 1b-e**). Next, we tested for associations between binarized patient characteristics and gut microbial community structure (**Supplemental Figure 1f**). Consistent with the Shannon diversity analysis, inter-individual differences in overall community structure were associated with prior antibiotic use (**Supplemental Figure 1f,g**).

### The human gut microbiome is altered after chemotherapy

To assess the impact of CAP treatment on the gut microbiome, we compared taxonomic and pathway abundance post-treatment (t_3_) relative to baseline (t_1_) (**Figure 1a**). After adjusting for multiple hypothesis testing, we identified 5 enriched and 5 depleted species (**Figure 1b**). While the overall trend was significant across the full cohort, further inspection revealed that these species were more dramatically affected in a subset of patients (**Figure 1c**). The 10 differentially abundant species were from similar higher-level taxonomic groups; multiple *Clostridiales* species were enriched, while multiple *Actinomycetaceae* species were depleted (**Figure 1d; Supplemental Table 2**). Pathway abundance was even more dramatically altered, with 257 significantly enriched and 2 significantly depleted pathways following CAP treatment (**Figure 1e, Supplemental Table 3**). Together, these findings reveal that despite the marked heterogeneity in patient characteristics and baseline microbial community structure, it is possible to identify consistent shifts in taxonomic composition and metabolic pathway abundance following three cycles of CAP treatment.

To investigate whether these microbiome shifts would be detected during treatment, we compared taxonomic and pathway abundance during cycle 3 (t_2_) relative to baseline (t_1_). The compositional differences were more modest at this earlier time with only two species reaching significance: *Slackia exigua* and *Clostridium sp. NSJ 42* (**Supplemental Figure 2a, Supplemental Table 2**). However, the overall trends were comparable, with a significant correlation in the fold-change of bacterial species relative to baseline, during cycle 3, and post-treatment (**Supplemental Figure 2b**). Similar trends were observed in the pathway analysis. A more modest set of pathways was significantly different during treatment, including 25 enriched and 3 depleted pathways (**Supplemental Figure 2c, Supplemental Table 3**). Nevertheless, there remained a significant correlation between pathway-level differences in relative abundance during and after treatment (**Supplemental Figure 2d**).

Notably, seven of the top ten most significantly enriched pathways post-treatment represented menaquinol biosynthesis or related pathways. Menaquinol is a reduced form of vitamin K2 (menaquinone) that is produced by diverse members of the gut microbiota and readily interconverted to menaquinone in bacterial and mammalian cells^38–40^. We investigated the microbial source of menaquinol biosynthesis using stratified pathway abundance data from our patient cohort. More than 70% of menaquinol biosynthesis abundance was attributable to *Escherichia* spp., with enrichment of *Escherichia* and unclassified sources responsible for the enrichment of menaquinol biosynthesis pathways following CAP treatment (**Figure 1f**). Next, we retrieved the KEGG orthologous groups (KOs) shared across all 7 enriched menaquinol biosynthesis pathways. All of these KOs were significantly enriched (**Figure 1g**). Analysis at a per-patient level revealed clear inter-individual differences in the temporal shifts in menaquinol biosynthesis pathway relative abundance, with 70.5% of patients exhibiting a net increase relative to baseline (**Figure 1h**). Patients who experienced menaquinol biosynthesis gene enrichment had significantly lower-stage disease at diagnosis (**Supplemental Figure 2e**), and were significantly more likely to require dose reductions during treatment (**Supplemental Figure 2f**).

### Menaquinol biosynthesis rescues bacterial fluoropyrimidine sensitivity

Because bacterial menaquinol biosynthesis genes were enriched following fluoropyrimidine treatment and are responsible for the production of menaquinones (vitamin K2)^39^, we hypothesized that menaquinol biosynthesis could be a protective factor allowing bacteria to escape the off-target effects of fluoropyrimidines on gut bacteria^13^. The model organism *E. coli* K-12 encodes an intact menaquinol biosynthesis pathway^41^, is sensitive to fluoropyrimidines^13^, and is genetically tractable^42^, providing a useful model system to test causal links between vitamin K2 production, anti-cancer drugs, and bacterial growth.

We leveraged a previously published genome-wide random barcode transposon-site sequencing (RB-TnSeq) library that covers 3,789 non-essential genes with a total of 152,018 unique transposon insertions^26^. The *E. coli* RB-TnSeq library was cultured for 48 hours in M9 with vehicle or 500 µM of three fluoropyrimidines that had all been previously shown to inhibit *E. coli* growth^13^: CAP, 5’-deoxy-5-fluorocytidine (DFCR), and 5-FU.

5-FU induced major overall changes in library composition (**Figure 2a, Supplemental Figure 3a,b, Supplemental Tables 4-6**). We identified a total of 513 protective (**Figure 2a**) and 274 detrimental (**Supplemental Figure 3b**) genes during incubation with any of the three fluoropyrimidines. A subset of genes were consistent across the three drugs, including 2 protective and 2 detrimental genes (**Supplemental Tables 4-6**). Transposon insertions in the uracil phosphoribosyltransferase (*upp*) gene were dramatically enriched in response to all three fluoropyrimidines (**Supplemental Figure 3c**), confirming its key role in exacerbating bacterial 5-FU toxicity^43^. On the other hand, dUMP phosphatase (*yjjG*) insertions were most dramatically depleted across conditions (**Figure 2b**), confirming its role in mitigating bacterial 5-FU toxicity by preventing incorporation of mutagenic nucleotides^44^.

**Figure 2.**
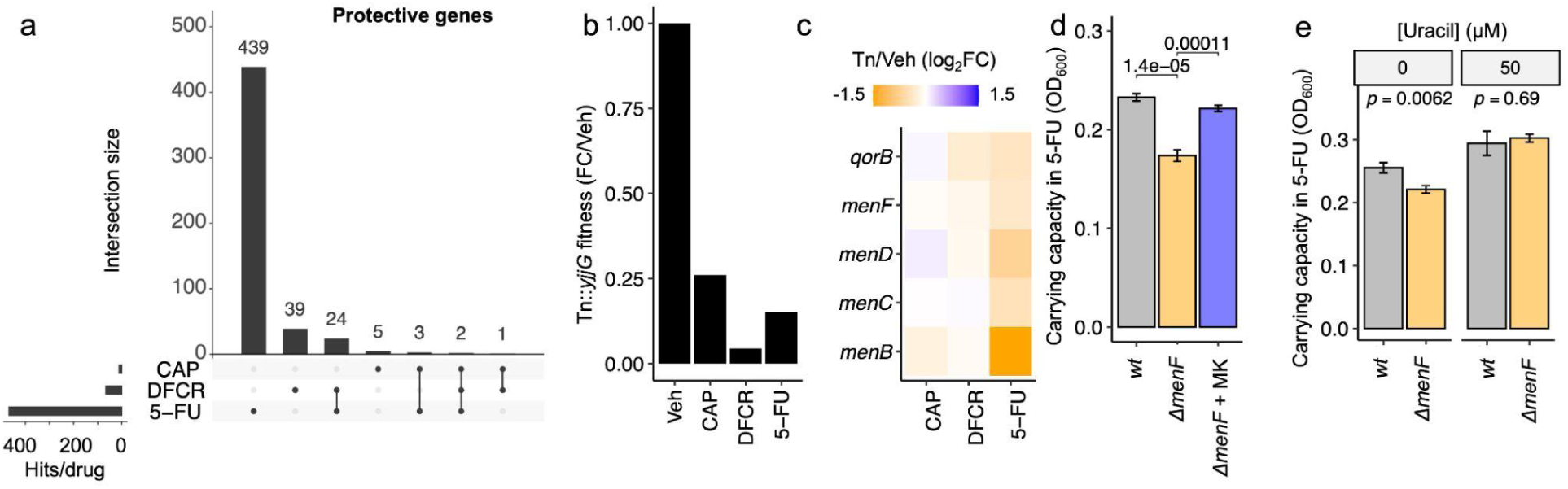
Menaquinol biosynthesis rescues bacterial sensitivity to fluoropyrimidines. **(a-c)** An *E. coli* RB-TnSeq library was treated with 500 μM of capecitabine (CAP), 5’-deoxy-5-fluorocytidine (DFCR), 5-fluorouracil (5-FU), or vehicle (Veh) in duplicate for 48 hours. **(a)** Upset plot of significantly depleted transposon-disrupted genes (intact gene is protective) across all 3 conditions. **(b)** Fitness of Tn::*yjjG* mutant in all four conditions, relative to vehicle. Values represent the mean of 2 biological replicates. **(c)** Gene set enrichment analysis of protective genes from **(a)** revealed quinone biosynthesis as the sole significantly enriched pathway (hypergeometric *p*<0.01). RB-TnSeq fold-change of enriched quinone biosynthesis genes is depicted. **(d-e)** *E. coli* BW25113 wild-type (*wt*) and *ΔmenF::Kan^R^* (*ΔmenF*) were treated with 500 μM 5-FU ± 225 nM menaquinone (MK) **(d)** or ± 50 μM uracil (**e**) for 24 hours, with carrying capacity quantified with Growthcurver. *p*-values: deviation from linearity on quantile-quantile plot **(a)**, Student’s t-test (**d,e**).

Next, we performed gene set enrichment analysis for genes that were enriched or depleted by at least one drug to gain a high-level view of the genetic determinants of fluoropyrimidine sensitivity. The detrimental genes in response to fluoropyrimidine treatment (5-FU, DFCR, and/or CAP) were significantly enriched for homologous recombination (*p*=0.0099, **Supplemental Figure 3d**), including transposon insertions in Holliday junction DNA helicase *ruvA/ruvB*, potentially due to enhanced cellular toxicity following inaccurate DNA damage repair. Protective genes in response to fluoropyrimidines were significantly enriched only for quinone biosynthesis (*p*=0.0056), including many of our previously identified genes for menaquinol biosynthesis (**Figure 1g**). Consistent with the broader pattern in this analysis, 5-FU led to a more marked depletion of menaquinol biosynthesis genes (**Figure 2c**).

Bacterial genetics and media supplementation validated a causal role of menaquinol biosynthesis in mediating protection from the off-target effects of fluoropyrimidines for bacterial growth. First, we acquired an in-frame, kanamycin (Kan) resistant single gene deletion of the first step of the menaquinol biosynthesis pathway (*menF*, K02552) from the Keio collection^29^. We grew *E. coli* BW25113 wild-type (*wt*) and Δ*menF::Kan^R^*in 0 and 500 μM 5-FU. Overall growth of the two strains was comparable in the absence of 5-FU (**Supplemental Table 7**). The carrying capacity of Δ*menF::Kan^R^* relative to *wt* decreased when subjected to 5-FU (**Figure 2d**). Next, we grew *E. coli* BW25113 Δ*menF::Kan^R^* in 5-FU with 0.1 μg/mL menaquinone. Menaquinone markedly rescued carrying capacity in the presence of 5-FU (**Figure 2d, Supplemental Table 7**).

Prior studies showed that menaquinol biosynthesis defects lead to uracil auxotrophy in *E. coli*^45–47^, suggesting this pathway may exert a chemoprotective effect via modulating uracil. To test whether uracil could rescue the 5-FU-dependent Δ*menF::Kan^R^* fitness defect, we grew *E. coli wt* and Δ*menF::Kan^R^* in 5-FU±50 μM uracil. While Δ*menF::Kan^R^* grew worse than *wt* in 5-FU in media, both strains grew comparably with uracil supplementation (**Figure 2e, Supplemental Table 7**). Taken together, these findings suggest that fluoropyrimidines directly select for bacteria with the ability to synthesize chemoprotective menaquinone, prompting us to consider the broader chemoprotective role of the microbiome in mediating host drug toxicities.

### Baseline gut microbial functional pathways are associated with toxicities in patients

Most patients experienced at least one patient-reported toxicity-related event (any grade) during treatment (*n*=45/48 patients with t_2_ toxicity data available; **Figure 3a**). To investigate whether the microbiome varies by toxicity status, we performed PERMANOVA testing comparing these on-treatment toxicities with the community composition of baseline species and pathway abundances. We did not find any significant relationships with baseline species abundance (FDR>0.2). In contrast, the composition of baseline pathway abundance was significantly associated with patient-reported PSN, alopecia, and oral mucositis (**Figure 3b**).

**Figure 3.**
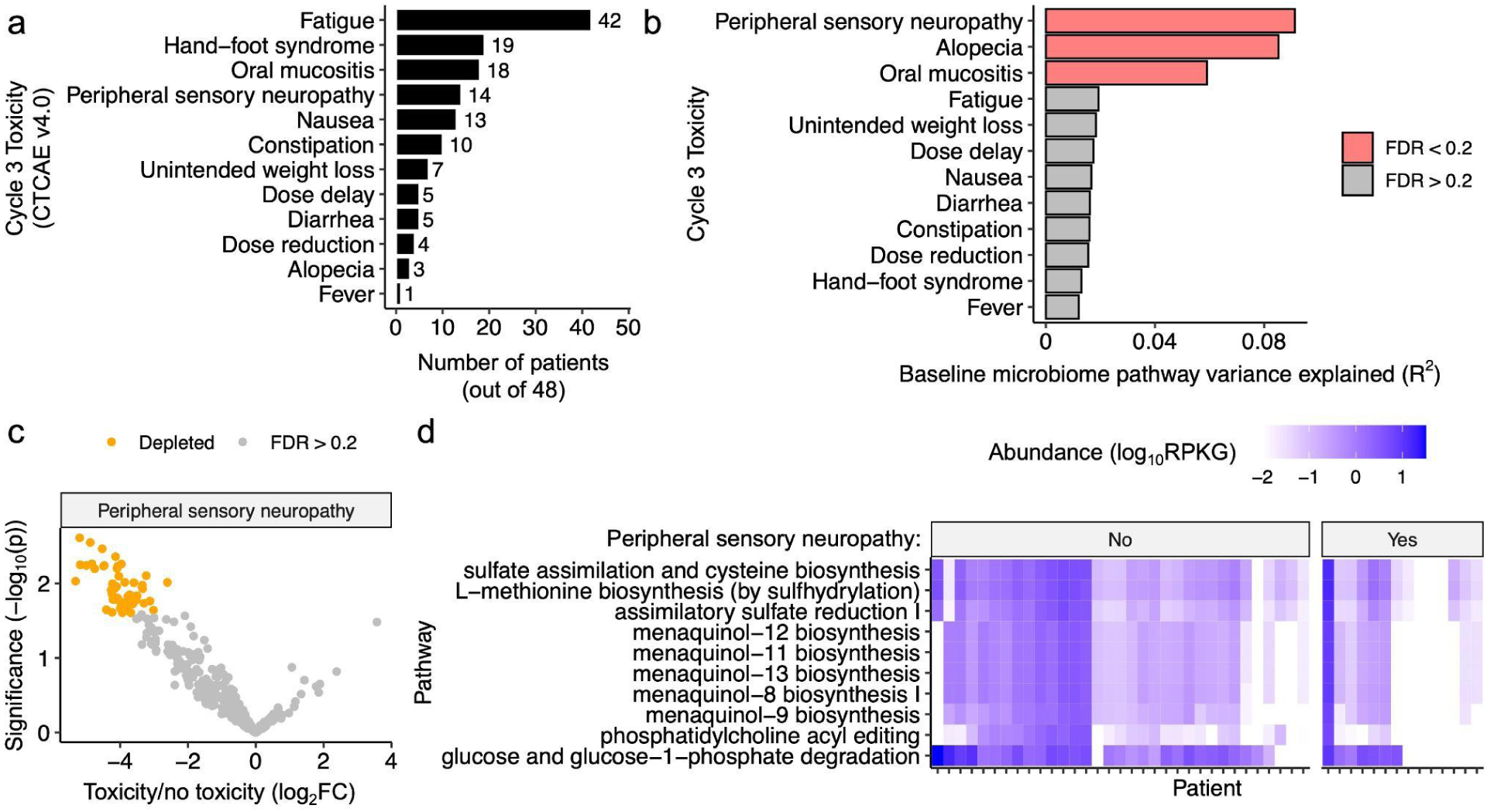
Pre-treatment microbial gene pathways are associated with development of toxicities during treatment. **(a)** Distribution of Grade 1+ toxicities in patients at cycle 3 (t_2_). **(b)** Permutational multivariate analysis of variance (PERMANOVA) testing of cycle 3 (t_2_) toxicities with respect to baseline bacterial gene family composition. *p*-value: PERMANOVA test using the central log ratio (CLR)-transformed Euclidean metric of baseline bacterial gene family composition, with FDR calculated with Benjamini-Hochberg multiple-testing correction. **(c)** Volcano plot of baseline gene pathways in patients who went on to have peripheral sensory neuropathy (PSN) or no PSN during treatment. Colored points represent significantly depleted (orange) pathways (FDR<0.2). *p*-value: linear model of abundance vs toxicity. **(d)** Heatmap of the baseline (t_1_) abundances of the top 10 lowest FDR pathways from **(c)** in units of reads per kilobase per genome equivalent (RPKG), faceted by whether a patient experienced PSN, with patients and pathways ordered by median hierarchical clustering.

Next, we compared baseline pathway abundance in patients who went on to report PSN-like symptoms, alopecia, and oral mucositis, and those who did not. We found a significant depletion in 59 pathways in patients who went on to experience PSN (**Figure 3c and Supplemental Table 8**). Remarkably, patients who reported PSN-like symptoms had significantly lower baseline levels of menaquinol biosynthesis pathways (**Figure 3d**), supporting the potential clinical relevance of our microbiome analyses (**Figure 1**) and experiments in bacterial cultures (**Figure 2**). Multiple pathways related to sulfate were also statistically significant (**Figure 3d**). Analysis of the cycle 3 and post-treatment data revealed that these pre-existing differences in PSN-associated pathways equalize in response to treatment (**Supplemental Figure 4a**).

Distinct pathways were observed for alopecia, with a significant depletion of 291 pathways at baseline in patients who experienced this alopecia (**Supplemental Figure 4b and Supplemental Table 9**). We did not observe a role for menaquinol biosynthesis pathways (**Supplemental Table 9**). Instead, we noted a depletion in pathways involved in L-methionine biosynthesis and β-(14)-mannan degradation (**Supplemental Figure 4c**). Oral mucositis was not associated with any individual pathways (FDR>0.2).

### Baseline gut microbial gene family abundance accurately predicts toxicity

We sought to build a model using the baseline microbiome to predict toxicity during CAP treatment. Rather than relying on gene pathways which encompass genes with broad functions, we turned to more granular KO abundance data. For each toxicity of interest, we used Boruta to select KOs of interest, trained a random forest model on this cohort (48 patients with on-treatment toxicity data available), and validated the model on an independent cohort (38 patients with on-treatment toxicity data available; **Figure 4a**). The validation cohort consisted of fluoropyrimidine-treated patients with CRC treated at University of California, San Francisco (ClinicalStudies.gov NCT04054908)^15^. Due to the availability of detailed HFS (any grade) and dose adjustment data in the validation clinical dataset, we opted to focus on these toxicity categories.

**Figure 4.**
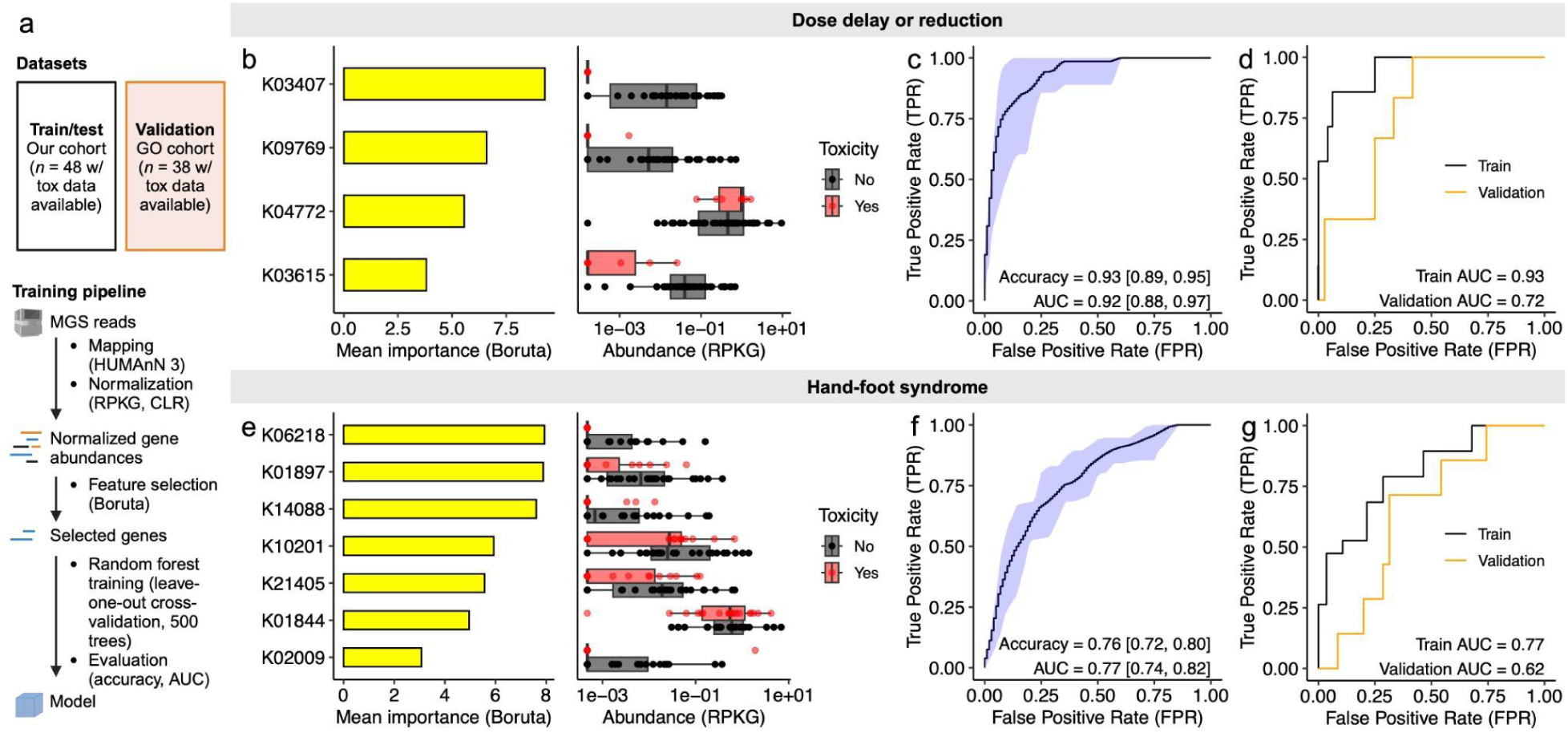
The baseline gut microbiome predicts drug side effect profiles. **(a)** Random forest pipeline. For each toxicity of interest, metagenomic sequencing reads were mapped to KEGG orthologous groups (KOs) using HUManN 3 and normalized by kilobase per genome equivalent (RPKG), followed by a central log ratio (CLR) transform, followed by feature selection with Boruta. A random forest algorithm was trained on these features using leave-one-out cross-validation (LOOCV) with 500 trees, followed by evaluation on our cohort and an independent validation cohort of 38 American patients with toxicity data available^15^. Created with BioRender.com. **(b,e)** Importance scores and baseline (t_1_) abundances of Boruta-selected KOs to classify dosing changes **(b)** or hand-foot syndrome (HFS) **(e)** during treatment (t_2_). **(c,f)** Receiver operating characteristic (ROC) curve for classification of dosing changes **(c)** or HFS **(f)** with random forest models built with Boruta-selected KOs tested with LOOCV. The black line represents the mean and blue shaded area represents the 95% confidence interval obtained across 10 independent models. Accuracy and area under the curve (AUC) are displayed, with 95% confidence intervals in brackets. **(d,g)** Evaluation of a model trained on our dataset and validated on an independent cohort of 38 American patients to predict dosing changes **(d)** or HFS **(g).**

In our training cohort, the Boruta algorithm selected four baseline microbial KOs as features relevant to the development of a model predicting dose adjustments (**Figure 4b**). Of these KOs, serine endoprotease *degQ* (K04772) was more abundant in dose-adjusted patients, while sensor kinase *cheA* (K03407), nucleotide metabolism esterase *ymdB* (K09769), and ion-translating oxidoreductase *rnfC* (K03615) were more abundant in patients who did not require dose delays or reductions (**Figure 4b**). Using these four genes, we trained 100 random forest models using leave-one-out cross-validation, achieving a mean accuracy of 0.93 and area under the curve (AUC) of 0.92 (**Figure 4c**). Finally, we validated a random forest model trained on our validation cohort, obtaining an AUC of 0.73 (**Figure 4d**).

For HFS, the Boruta algorithm selected seven baseline microbial KOs as relevant features in our training cohort (**Figure 4e**). All of these KOs were less abundant in patients who experienced HFS (**Figure 4e**). Using these 7 genes, we trained 100 random forest models using leave-one-out cross-validation, achieving a mean accuracy of 0.76 and area under the curve (AUC) of 0.77 (**Figure 4f**). The validation cohort AUC was 0.62 (**Figure 4g**).

Finally, inspired by our observations associating baseline pathway abundance and PSN, we sought to develop an algorithm to predict patient-reported PSN in our cohort in spite of a lack of validation cohort. We selected the top 10 differentially abundant pathways (**Figure 3c,d**) and trained 100 random forest models using leave-one-out cross-validation, achieving a mean accuracy of 0.77 and area under the curve (AUC) of 0.72 (**Supplemental Figure 5**).

## Discussion

Our metagenomic and experimental data revealed an unexpected role for microbial vitamin K2 biosynthesis in protection from the off-target effects of fluoropyrimidines on gut bacterial growth. The primary mechanism-of-action of 5-FU, thymidylate synthase inhibition, does not have any direct links to vitamin K2, in contrast to other micronutrients including folate and vitamin B6^48, 49^. Our experiments in *E. coli* suggest this pathway may exert a chemoprotective effect via modulating uracil, protecting bacteria from 5-FU^13^.

These data support a protective role of microbial menaquinol biosynthesis in ameliorating aspects of host drug toxicity. Higher baseline levels of menaquinol biosynthesis genes were associated with decreased risk of patient-reported PSN. Consistent with this data, demyelination of peripheral nerve fibers is a primary cause of PSN^50^ and vitamin K2 plays a crucial role in the myelin sheath repair in the peripheral nervous system^51^. Vitamin K2 supplementation can alleviate peripheral neuropathies in patients with vitamin B12 deficiency or type 2 diabetes mellitus^52^.

More broadly, we identified microbial biomarkers of drug toxicity across multiple endpoints (PSN, alopecia, oral mucositis). We found lower levels of mannan degradation genes in the gut microbiomes of subjects who developed alopecia. Intraperitoneal mannan delivery induces alopecia in a mouse model^53^. Thus, the balance between mannan consumption through diet, fungal production within the gastrointestinal tract, and gut bacterial degradation may modulate alopecia through systemic mannan levels.

Our data provides a *proof-of-concept* for the development of microbiome-based machine learning models that accurately predict drug toxicity in cancer chemotherapy patients, building upon prior studies in rheumatoid arthritis, prostate cancer radiotherapy, and immune checkpoint inhibitor-induced colitis^54–57^. Remarkably, these models required just a handful of KOs (4-7), which could be measured using less expensive targeted assays like quantitative PCR. A critical next step will be designing larger intervention studies to test the utility of such models in clinical decision making.

The current dataset has multiple limitations to address in subsequent efforts. We did not collect samples during the first two treatment cycles, potentially missing dramatic early-treatment shifts observed in a cohort of US CRC patients^15^. While our sample size (56 subjects, 156 samples) was sufficient to reach statistical significance and uncover interesting biology, it remains insufficient to inform concrete patient care guidelines. The observational nature of our study and lack of dietary data makes causal inferences challenging, a limitation partially overcome by our experimental validation.

Many patients in this cohort reported PSN-like symptoms, a toxicity more commonly associated with oxaliplatin than with capecitabine. The reported PSN-like symptoms may be related to HFS, a dose-limiting toxicity of capecitabine, which initially manifests with dysesthesia and tingling similar to neuropathy. Alternatively, capecitabine might worsen subclinical PSN in patients previously treated with oxaliplatin^58^. In our cohort, 10/14 patients reporting PSN had previous oxaliplatin exposure (**Supplemental Table 1**). Although PSN is less common than HFS, previous studies have also observed PSN in 16-37% of patients treated with capecitabine or 5-FU without oxaliplatin^12, 59^.

In conclusion, our findings provide further support for a role of the gut microbiome in mediating the cancer treatment outcomes and the utility of paired studies in well-characterized patient cohorts and experimental model systems. These results raise numerous testable hypotheses that should be explored in preclinical models. Future work should focus on controlled clinical intervention studies to investigate if the use of vitamin supplementation, probiotics, or other microbiome-based interventions can alleviate drug toxicity in cancer patients.

## Data Availability

All raw sequencing data with human reads removed have been deposited to the NCBI Sequence Read Archive (SRA) and are accessible via BioProject PRJNA1169175. Processed datasets and all original code used in this study is available on GitHub (https://github.com/turnbaughlab/2024_Trepka_DrugToxicity).

## Acknowledgements

Sequencing was performed at Chan-Zuckerberg Biohub-San Francisco. Funding was provided from the National Institutes of Health (R01CA255116, R01DK114034, and R01HL122593 to P.J.T.) and Stichting Jules Coenegracht Sr (to M.L.S.). P.J.T is a Chan Zuckerberg Biohub-San Francisco Investigator. Diagrams were created with Biorender.com.

## Author contributions

**Lars E. Hillege**: Conceptualization, Methodology, Validation, Investigation, Data curation, Writing – Original draft, Writing – Review & Editing

**Kai R. Trepka**: Conceptualization, Methodology, Software, Validation, Formal Analysis, Investigation, Writing – Original draft, Writing – Review & Editing, Visualization

**Janine Ziemons**: Conceptualization, Methodology, Validation, Investigation, Data curation, Writing – Original draft, Writing – Review & Editing

**Romy Aarnoutse**: Conceptualization, Methodology, Investigation, Writing – Review & Editing, Funding acquisition

**Benjamin G. H. Guthrie**: Methodology, Investigation

**Judith de Vos-Geelen**: Conceptualization, Methodology, Resources, Data curation, Writing – Review & Editing, Supervision

**Liselot Valkenburg-van Iersel**: Resources, Data curation, Writing – Review & Editing

**Irene E.G. van Hellemond**: Resources, Data curation, Writing – Review & Editing

**Arnold Baars**: Resources, Data curation, Writing – Review & Editing

**Johanna H.M.J. Vestjens**: Resources, Data curation, Writing – Review & Editing

**John Penders**: Conceptualization, Methodology, Validation, Recourses, Writing – Review & Editing, Supervision

**Adam Deutschbauer**: Methodology, Resources

**Chloe E. Atreya**: Writing – Review & Editing, Supervision

**Wesley A. Kidder**: Writing – Review & Editing, Supervision

**Peter J. Turnbaugh**: Conceptualization, Methodology, Writing – Review & Editing, Supervision, Funding acquisition

**Marjolein L. Smidt**: Conceptualization, Methodology, Validation, Writing – Review & Editing, Supervision, Funding acquisition

## Supplemental Figures and Figure Legends

**Supplemental Figure 1.**
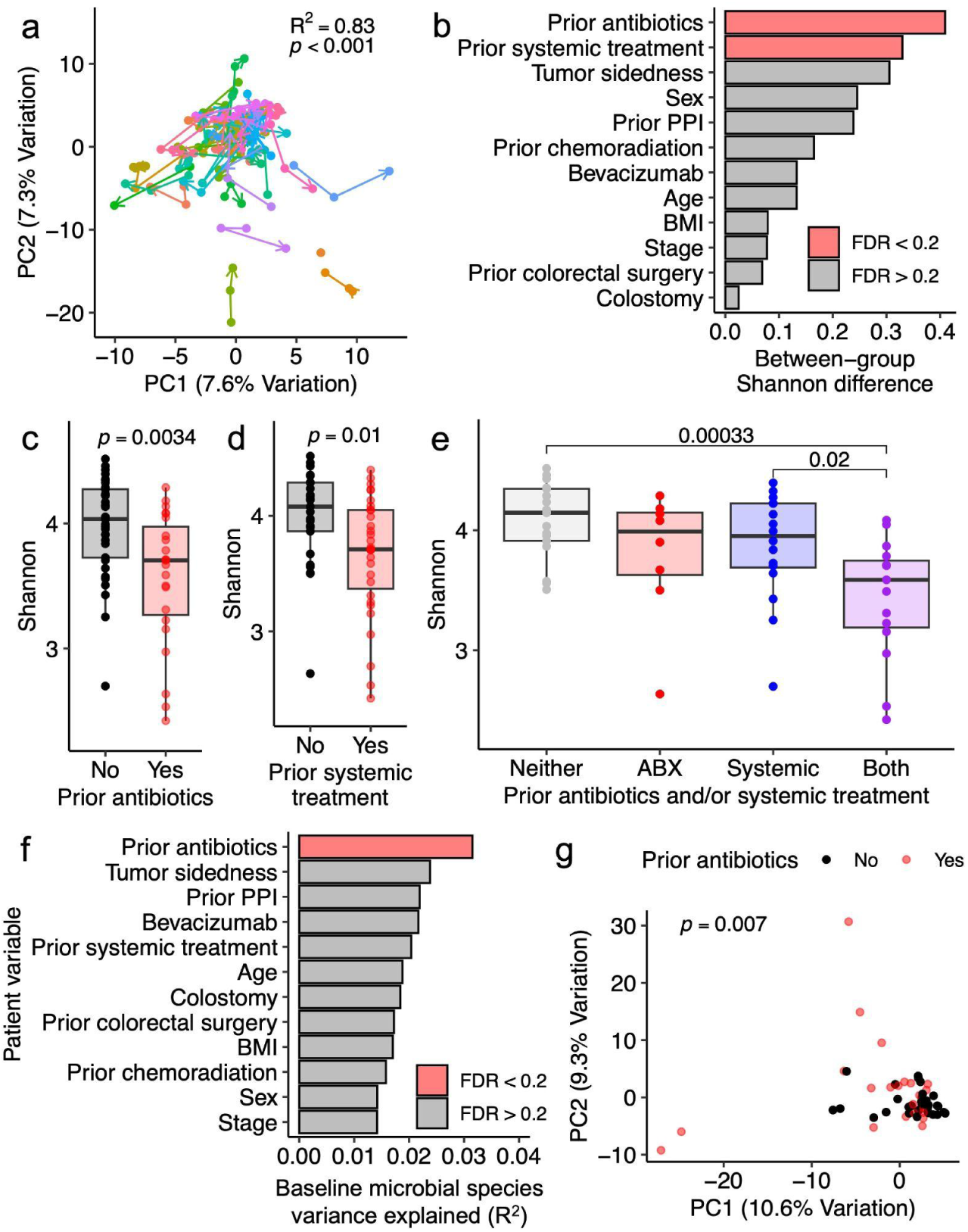
Prior antibiotic use and systemic treatment are associated with altered baseline microbial diversity. **(a)** Species-level principal coordinates analysis (central log ratio (CLR)-transformed Euclidean distances) across all timepoints, colored by patient, with arrows connecting patient samples pointing towards later timepoints (i.e baseline, cycle 3, post). **(b)** Patient demographics and treatment history are associated with bacterial diversity differences (Shannon index) at baseline. **(c-e)** Boxplots of Shannon diversity vs antibiotic use **(c)**, prior systemic treatment **(d)**, or antibiotic use and/or prior systemic treatment (**e**). **(f)** Permutational multivariate analysis of variance (PERMANOVA) testing of patient demographics and treatment history with respect to baseline bacterial taxa composition. **(g)** PCA of CLR-transformed Euclidean distances depicting antibiotic-associated differences in the baseline microbial species. *p-*values: Student’s *t* test **(b-e)**, PERMANOVA test with central log ratio (CLR)-Euclidean ordination **(a,f,g).** For **(c-e)**, comparisons with *p*<0.05 are labeled. Benjamini-Hochberg false discovery rate (FDR) correction applied for **(b,f)**, with FDR<0.2 called as significant.

**Supplemental Figure 2.**
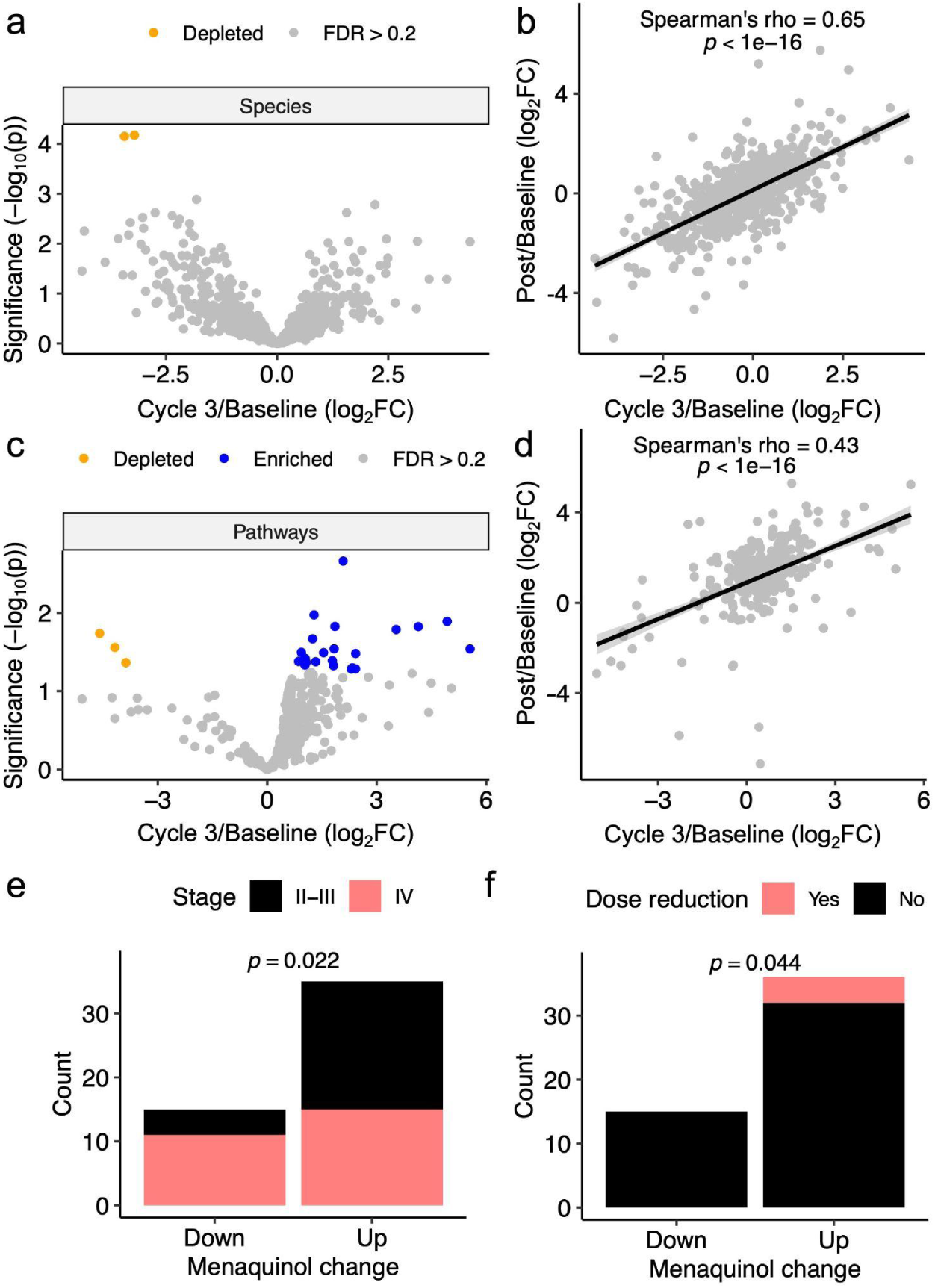
Consistent microbiota shifts during and after treatment. **(a)** Volcano plot of differentially abundant microbial species during cycle 3 (t_2_) vs baseline (t_1_). Orange dots represent significantly depleted species [false discovery rate (FDR)<0.2]. **(b)** Comparison of log_2_ fold change of species at cycle 3 (t_2_) or post-treatment (t_3_) relative to baseline (t_1_). **(c)** Volcano plot of pathways during cycle 3 (t_2_) vs baseline (t_1_). Points represent significantly enriched (blue) and depleted (orange) pathways (FDR<0.2). **(d)** Comparison of log2 fold change of pathways at cycle 3 (t_2_) or post-treatment (t_3_) relative to baseline (t_1_). **(e,f)** Comparison of menaquinol synthesis gene enrichment during treatment versus cancer stage (**e**) and on-treatment (t_2_) dose reduction (**f**). Patients were grouped using Average log_2_FC depicted in **Figure 1h**. *p*-values: Mixed-effects model of abundance vs time, with patient as a random effect (**a,c**); Spearman’s rank correlation (**b,d**); one-sided likelihood-ratio test (**e,f**).

**Supplemental Figure 3.**
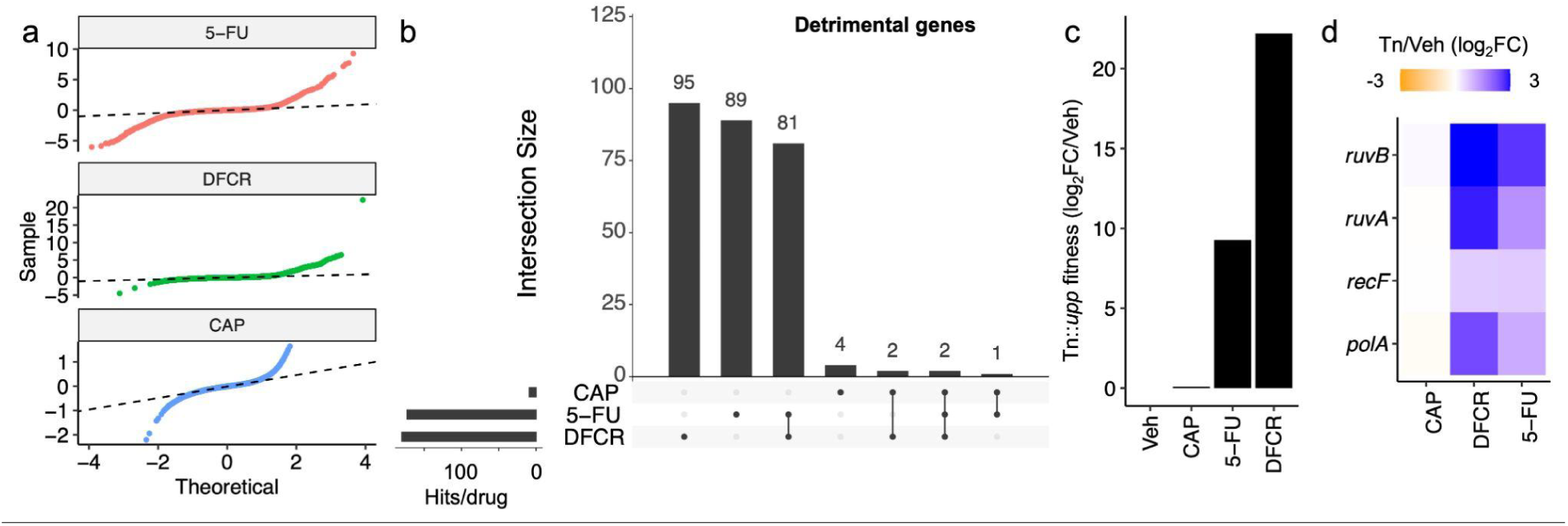
Uridine phosphorylase (*upp*) exacerbates fluoropyrimidine toxicity. **(a)** Quantile-quantile (Q-Q) plot showing deviation from normality (dotted black line) for a RB-TnSeq library treated with 500 μM capecitabine (CAP), 5’deoxy-5-fluorocytidine (DFCR), and 5-fluorouracil (5-FU) relative to Vehicle (Veh). **(b)** Upset plot of significantly enriched transposon-disrupted genes (i.e. the intact gene is detrimental) across all 3 conditions. **(c)** Fitness of Tn::*upp* mutant in all four conditions, relative to vehicle. Values represent the mean of 2 biological replicates. **(d)** Gene set enrichment analysis of detrimental genes from **(b)** revealed homologous recombination as the sole significantly enriched pathway (*p*<0.01). RB-TnSeq fold-change of enriched quinone biosynthesis genes is depicted.

**Supplemental Figure 4.**
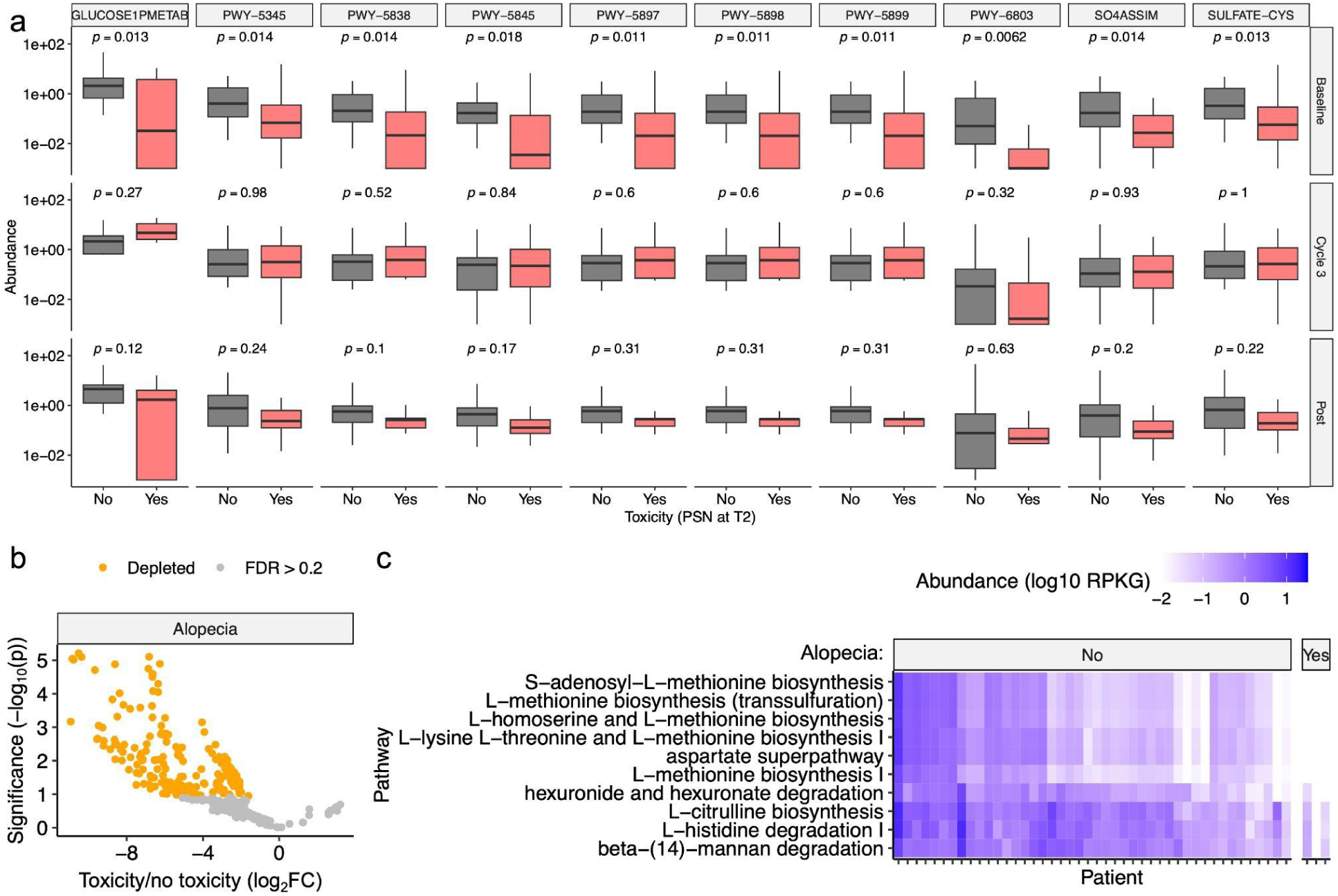
Pretreatment microbial gene pathways are associated with development of toxicities during treatment. **(a)** Abundance of top 10 pathways significantly associated with peripheral sensory neuropathy (PSN) at t_2_ (pathways from **Fig. 3d**, labeled with MetaCyc pathway numbers), faceted by pathway and time of stool sample. *p*-values: ANOVA. **(b)** Volcano plot of baseline gene pathways in patients who went on to have alopecia or no alopecia during treatment. Colored points represent significantly depleted (orange) pathways (FDR<0.2). *p*-value: linear model of abundance vs toxicity. **(c)** Heatmap of the baseline (t_1_) abundances of the top 10 lowest FDR pathways from **(b)** in units of reads per kilobase per genome equivalent (RPKG), faceted by whether a patient experienced alopecia, with patients and pathways ordered by median hierarchical clustering.

**Supplemental Figure 5.**
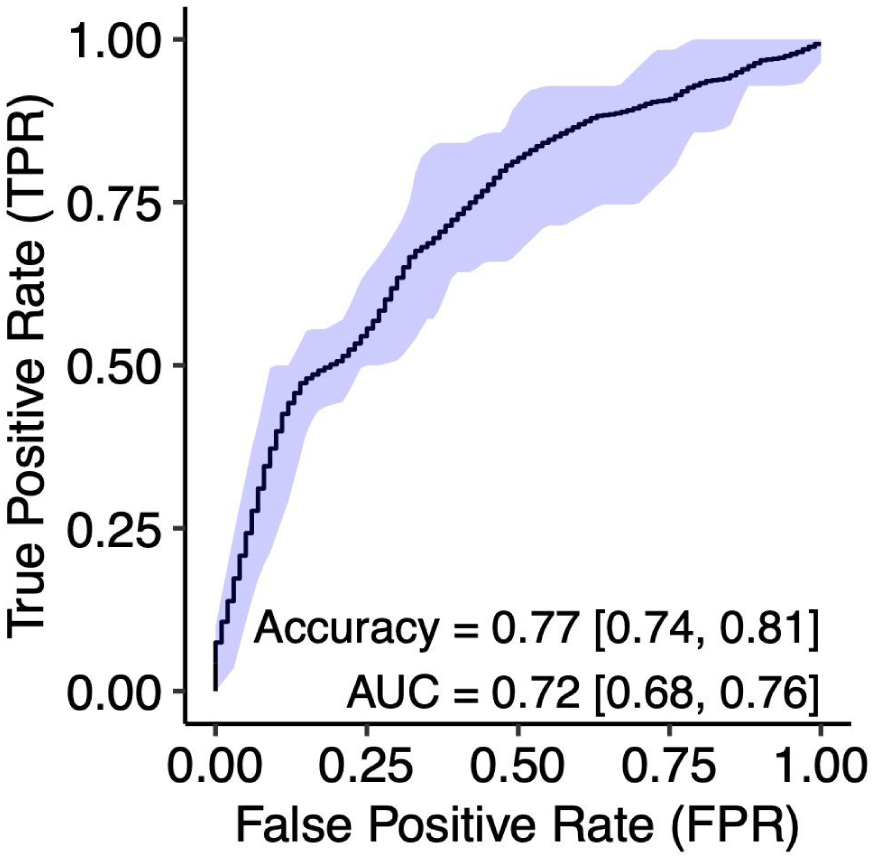
Pretreatment microbial gene pathways predict peripheral sensory neuropathy during treatment (at t_2_). Receiver operating characteristic (ROC) curve for classification of tumor change with random forest models built with pathways identified in **Fig. 3d**, tested with leave-one-out cross-validation. The black line represents the mean and blue shaded area represents the 95% confidence interval obtained across 100 independent models. Accuracy and area under the curve (AUC) are displayed, with 95% confidence intervals in brackets.

